# Substantial SF-36 score differences according to the mode of administration of the questionnaire: an ancillary study of the SENTIPAT multicenter randomized controlled trial comparing web-based questionnaire self-completion and telephone interview

**DOI:** 10.1101/2021.02.08.21251357

**Authors:** Ayşe Açma, Fabrice Carrat, Gilles Hejblum, for the SENTIPAT study group

## Abstract

**Background:** SF-36 is a popular questionnaire for measuring self-perception of quality of life in a given population of interest. Surprisingly, no study compared score values issued from a telephone interview versus an internet-based questionnaire self-completion.

**Methods:** Patients having an Internet connection and returning home after hospital discharge were enrolled in the SENTIPAT multicenter randomized trial the day of discharge. They were randomized to either self-complete a set of questionnaires using a dedicated website (I group) or to provide answers to the same questionnaires administered during a telephone interview (T group). This ancillary study of the trial compared SF-36 data relating to the post-hospitalization period in these two groups. In order to anticipate potential unbalanced characteristics of the respondents in the two groups, the impact of the mode of administration of the questionnaire on score differences was investigated using a matched sample of individuals originating from I and T groups (ratio 1:1), the matching procedure being based on a propensity score approach. SF-36 scores observed in I and T groups were compared with a Wilcoxon-Mann-Whitney test, the score differences between the two groups were also examined according to Cohen’s effect size.

**Results:** There were 245/840 (29%) and 630/840 (75%) SF-36 questionnaires completed in the I and T group, respectively (p < 0.001). Globally, score differences between groups before matching were similar to those observed in the matched sample. Mean scores observed in T group were all above the corresponding values observed in the I group. After matching, score differences in six out of the eight SF-36 scales were statistically significant, with a mean difference greater than 5 for four scales and an associated mild effect size ranging from 0.22 to 0.29, and with a mean difference near this threshold for two other scales (4.57 and 4.56) and a low corresponding effect size (0.18 and 0.16, respectively).

**Conclusions:** Telephone mode of administration of SF-36 involved an interviewer effect increasing SF-36 scores. Questionnaire self-completion via the Internet should be preferred and surveys combining various administration methods should be avoided.

**Trial Registration:** ClinicalTrials.gov NCT01769261, registered January 16, 2013.

## Background

A query exploring the presence of the term “SF-36” in the title or the abstract of PubMed records retrieved 21058 documents on December 16, 2020: developed at RAND Corporation as part of the Medical Outcomes Study, the 36-Item Short Form Health Survey (SF-36) is indeed a popular questionnaire for measuring self-perception of quality of life (QoL) in a given population of interest [1-3]. SF-36 has been made available in 50 different languages including French [4]. While SF-36 was initially developed as a paper-pencil format auto-questionnaire, use of telephone interviews has been also reported for collecting SF-36 data [5-8]. Self-completion via the Internet has been reported as a validated administration mode by Bell and Kahn in 1996 [9] and since then, with the spread of wide internet and computers, several other computerized or internet based formats have been applied in different studies [10-12].

Several randomized trials compared the SF-36 scores issued from different administration modes, such as paper versus internet [13-17] or telephone versus paper [18-26]. Telephone interview is a common mode of questionnaire administration for several reasons, including the potential to increase response rate [24, 26], practical convenience if other data of the study are already being collected via telephone, and exploring quality of life in some special populations such as very elderly patients. On the other hand, self-completion via the Internet has advantages like avoiding any potential response bias related to interviewer effect [18], being potentially a simpler organization for collecting SF-36 data, and associated with lower costs. However, and surprisingly, to our knowledge, no study compared telephone interview versus internet-based auto-questionnaire methods for collecting SF-36 data to investigate whether they can be used as alternative methods in the mixed-mode data collection procedures according to participant preferences and/or to minimize the possible selection bias.

In this context, the multicenter SENTIPAT (the concept of sentinel patients who would voluntarily report their health evolution on a dedicated website) randomized trial [27-29] is the first multicenter randomized trial comparing the Internet against telephone interviews as the methods of administrating several questionnaires on the health evolution of hospitalized patients. The aim of the present work is to compare SF-36 data relating to the 6-weeks post-discharge period of hospitalized patients, collected either via the Internet or through telephone interviews in the SENTIPAT trial.

## Methods

This research was an ancillary study of the multicenter, randomized SENTIPAT trial [27]. We took advantage of the trial to analyze patients’ QoL during the post-hospitalization period.

### Population

Briefly, as previously reported [28, 29], subjects recruited consecutively from 5 different volunteered units (Hepato-Gastroenterology, Gastrointestinal Enterology and Nutrition, General and Digestive Surgery, Infectious and Tropical Diseases, Internal Medicine) of Hôpital Saint Antoine were enrolled in the SENTIPAT trial. Patients with internet access at home, aged 18 or above, not cognitively impaired and without a behavioral disorder, speaking French, returning home after hospitalization, and not opposed to participating to the trial were eligible for inclusion. Eligible patients not opposed to participate in the study were randomized into two parallel groups: Internet (I) or telephone (T) follow-up (inherently resulting in an open-label trial) at a ratio of 1:1.

Inpatients were enrolled on the day of hospital discharge by a clinical research technician of the trial. At that time, patients were informed about the study. Eligible patients not opposed to participate in the study were randomized into two parallel groups: Internet or telephone follow-up (inherently resulting in an open-label trial) at a ratio of 1:1. On the basis of a centralized randomization that allocated the eligible patient either to the Internet or to the telephone group through a website and using an underlying permutation block randomization stratified by service, the computer-generated list of permutation was established by a statistician independent from the study. At the time of patient inclusion, the technician also collected baseline variables (length of stay, sex, age, relationship status, level of education, activity, and type of insurance). Patient was then informed and discharged with documents explaining corresponding questionnaire administration. A total of 1680 eligible patients (840 randomized in the I group and T group each) were enrolled in the SENTIPAT trial between February 25, 2013 and September 8, 2014.

### Survey administration

Patients of the I group had access to the SF-36 questionnaire 40 days after discharge on a web site dedicated to SENTIPAT. Patients of the T group were interviewed by telephone approximately 42 days after discharge and the data entered to a similar web site interface as used in the I group. In case of nonresponse, reminding emails were sent in the I group while up to three calls were tried whenever the first call did not reach the patient in the T group.

### SF-36 score calculations

The eight scale scores and the two summary scores of SF-36 were calculated according to MOS SF-36 French scoring manual [30]. The scale score calculations were done for the multi-item scales only if at least half of the items were answered and the missing item data, if existed, were treated with a person-specific approach which uses the average score of the completed items in the same scale.

### Statistical analyses

Bivariate analyses were performed using Fisher exact test or Chi-Square test of independence for the categorical variables, and the Wilcoxon-Mann-Whitney test for the quantitative variables. The latter test was notably used for comparing the SF-36 score differences between I and T groups. Several authors have discussed the task of interpreting observed differences in terms of “clinically meaningful” differences [31-33]. In this study, in addition to the above-mentioned statistical test, SF-36 score differences between I and T groups were also examined at the light of two popular approaches: on the one hand, effect size of the difference was considered according to Cohen’s effect size index [34]; on the other hand, we considered a threshold difference of five points, as was proposed by Ware et al [33] for defining a clinically and socially relevant difference between two compared scores. Internal reliability of the SF-36 was evaluated by Cronbach’s alpha coefficient calculation for the eight scales, and was considered as acceptable if the alpha value was > 0.7.

To determine the reasons for the potential score differences between the groups, a responder sample was composed of internet responders matched to telephone responders according to a propensity score-based procedure : the R package MatchIt [35] was used for matching each internet responder to the nearest telephone responder with a 1:1 ratio, and we forced each pair to be strictly identical according to three qualitative variables, sex (male / female), type of hospitalization (conventional / weekly / day-care hospitalization), and hospital ward (general and digestive surgery / gastroenterology and nutrition / hepato-gastroenterology / infectious and tropical diseases / internal medicine). The following baseline variables were additionally included in the logistic regression modeling the propensity score (propensity for being an internet responder versus being a telephone responder): age, length of stay, education, employment (unemployed because of health / retired or unemployed / job-seeker, employed, student), income, relationship status, and type of health insurance.

## Results

Figure 1 presents the flowchart of the study and Table 1 indicates baseline characteristics of the patients who constituted the population investigated in this study. The response rate observed in the I group (245/840, 29%) was significantly lower (p < 0.001) than that observed in the T group (630/840, 75%). The median [interquartile range] delay between hospital discharge and questionnaire completion was 42 [40; 46] and 42 [42; 46] days in responders of the I and T group, respectively.

**Table 1:** Demographic characteristics of responders and nonresponders in the Internet and Telephone groups.

| Feature | Internet |  | Telephone |  |
| --- | --- | --- | --- | --- |
|  | Responders, n = 245 | Nonresponders, n = 595 | Responders, n = 630 | Nonresponders, n = 210 |
| **Sex** |  |  |  |  |
| Female | 109(44.5)<sup>a</sup> | 269(45.2) | 254(40.3) | 103(49.0) |
| Male | 136(55.5) | 326(54.8) | 376(59.7) | 107(51.0) |
| **Age: mean, median[IQR] (years)** | 49.5; 50[37–61] | 46.6; 47[33–59] | 47.2; 47[34–58] | 43.8; 41[30–54] |
| **Length of stay: mean, median[IQR]** | 4.0; 1[1–5] | 4.0; 1[1–5] | 4.0; 1[1–5] | 4.1; 1[1–6] |
| **Type of hospitalization** |  |  |  |  |
| Conventional | 102(41.6) | 256(43.0) | 269(42.7) | 91(43.3) |
| One-day stay | 120(49.0) | 285(47.9) | 297(47.1) | 103(49.1) |
| Week stay | 23(9.4) | 54(9.1) | 64(10.2) | 16(7.6) |
| **Ward** |  |  |  |  |
| General and digestive surgery | 67(27.3) | 138(23.2) | 161(25.6) | 44(21.0) |
| Gastroenterology and Nutrition | 65(26.5) | 139(23.4) | 147(23.3) | 59(28.1) |
| Hepato-Gastroenterology | 28(11.4) | 75(12.6) | 75(11.9) | 29(13.8) |
| Infectious and Tropical Diseases | 55(22.4) | 150(25.2) | 160(25.4) | 45(21.4) |
| Internal Medicine | 30(12.2) | 93(15.6) | 87(13.8) | 33(15.7) |
| **Employment** |  |  |  |  |
| Currently employed | 158(65.0) | 353(59.3) | 375(59.5) | 132(63.2) |
| Job-seeker | 17(7.0) | 43(7.2) | 47(7.5) | 15(7.2) |
| Retired | 47(19.3) | 98(16.5) | 101(16.0) | 29(13.9) |
| Student | 6(2.5) | 38(6.4) | 48(7.6) | 17(8.1) |
| Doesn't work because of health | 11(4.5) | 48(8.1) | 49(7.8) | 11(5.3) |
| Without work | 2(0.8) | 9(1.5) | 8(1.3) | 4(1.9) |
| Other | 2(0.8) | 6(1.0) | 2(0.3) | 1(0.5) |
| **Type of employment** |  |  |  |  |
| Farmer | 0(0.0) | 1(0.0) | 0(0.0) | 0(0.0) |
| Self-employed, trader | 4(1.6) | 25(4.2) | 27(4.3) | 11(5.3) |
| Manager | 80(32.7) | 135(22.7) | 159(25.2) | 49(23.4) |
| Intermediate Profession | 39(15.9) | 91(15.3) | 105(16.7) | 31(14.8) |
| Middle-class occupation | 52(21.2) | 135(22.7) | 123(19.5) | 55(26.3) |
| Employee | 5(2) | 20(3.4) | 25(4) | 8(3.8) |
| Worker | 42(17.1) | 83(13.9) | 92(14.6) | 22(10.5) |
| No work | 23(9.4) | 105(17.6) | 99(15.7) | 33(15.8) |

**Table 1 (continued): Demographic characteristics of responders and nonresponders in the Internet and Telephone groups**
| Feature | Internet |  | Telephone |  |
| --- | --- | --- | --- | --- |
|  | Responders, n = 245 | Nonresponders, n = 595 | Responders, n = 630 | Nonresponders, n = 210 |
| **Level of education** |  |  |  |  |
| Primary or less | 18(7.3) | 58(9.7) | 47(7.5) | 31(14.8) |
| High school | 75(30.6) | 193(32.4) | 178(28.3) | 60(28.7) |
| Superior short-time | 37(15.1) | 95(16.0) | 94(14.9) | 33(15.8) |
| Graduate or post graduate | 115(46.9) | 249(41.8) | 311(49.4) | 85(40.7) |
| **Relationship status** |  |  |  |  |
| Living alone<sup>b</sup> | 103(42.0) | 291(48.9) | 293(46.5) | 121(57.9) |
| Living as a couple<sup>c</sup> | 142(58.0) | 304(51.1) | 337(53.5) | 88(42.1) |
| **Income level** |  |  |  |  |
| <450€ | 6(2.4) | 28(4.7) | 31(4.9) | 10(4.8) |
| [450€–1000€[ | 3(1.2) | 37(6.2) | 31(4.9) | 11(5.3) |
| [1000€–1500€[ | 17(6.9) | 61(10.3) | 51(8.1) | 17(8.1) |
| [1500€–2100€[ | 34(13.9) | 75(12.6) | 78(12.4) | 27(12.9) |
| [2100€–2800€[ | 26(10.6) | 70(11.8) | 66(10.5) | 25(12.0) |
| [2800€–4200€[ | 44(18.0) | 79(13.3) | 108(17.1) | 28(13.4) |
| ≥4200€ | 43(17.6) | 64(10.8) | 82(13.0) | 16(7.7) |
| No response | 72(29.4) | 181(30.4) | 183(29.0) | 75(35.9) |
| **Type of employment** |  |  |  |  |
| State medical help or universal health | 2(0.8) | 26(4.4) | 24(3.8) | 8(3.8) |
| Compulsory health insurance | 15(6.1) | 43(7.2) | 43(6.8) | 26(12.4) |
| Compulsory health insurance plus<br>complementary private health<br>insurance | 228(93.1) | 526(88.4) | 563(89.4) | 175(83.7) |

<sup>a</sup>All data of the Table are expressed as n(%), unless otherwise indicated.
<sup>b</sup>Single, widowed, divorced, separated.
<sup>c</sup>Married, living together under a civil solidarity pact, simply living together without legal ties.
Abbreviation used: IQR, interquartile range.

**Figure 1.**
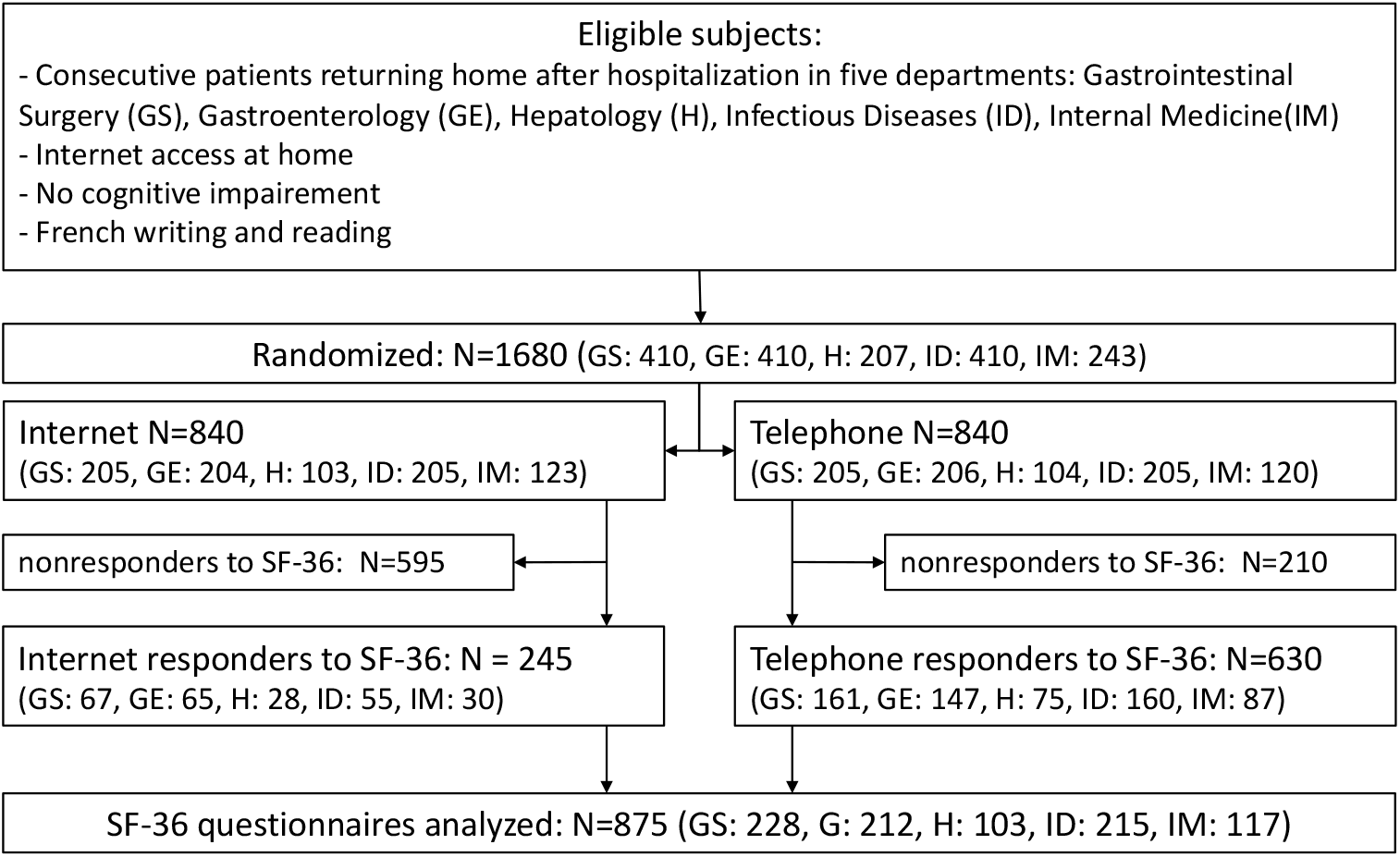
Flow of participants through the study.

In terms of internal validity of questionnaire completion, Cronbach’s alpha values calculated for each of the eight scales composing the SF–36 form in the I and T groups (see Supplemental Table) were all > 0.7, the threshold value considered as acceptable. The matching procedure matched the 245 respondents of group I–no individual was dropped–with 245 individuals of group T. The standardized mean difference of the global distance between I and T was 0.4167 and 0.0215 before and after matching, respectively, with a corresponding balance improvement of 95%. Figure 2 details the standardized mean differences between I and T groups observed on baseline variables, before and after the matching procedure. The differences between I and T groups before matching were globally dramatically dropped after matching, indicating that the matching procedure successfully yielded two populations I and T highly comparable in terms of the baseline variables.

**Figure 2:**
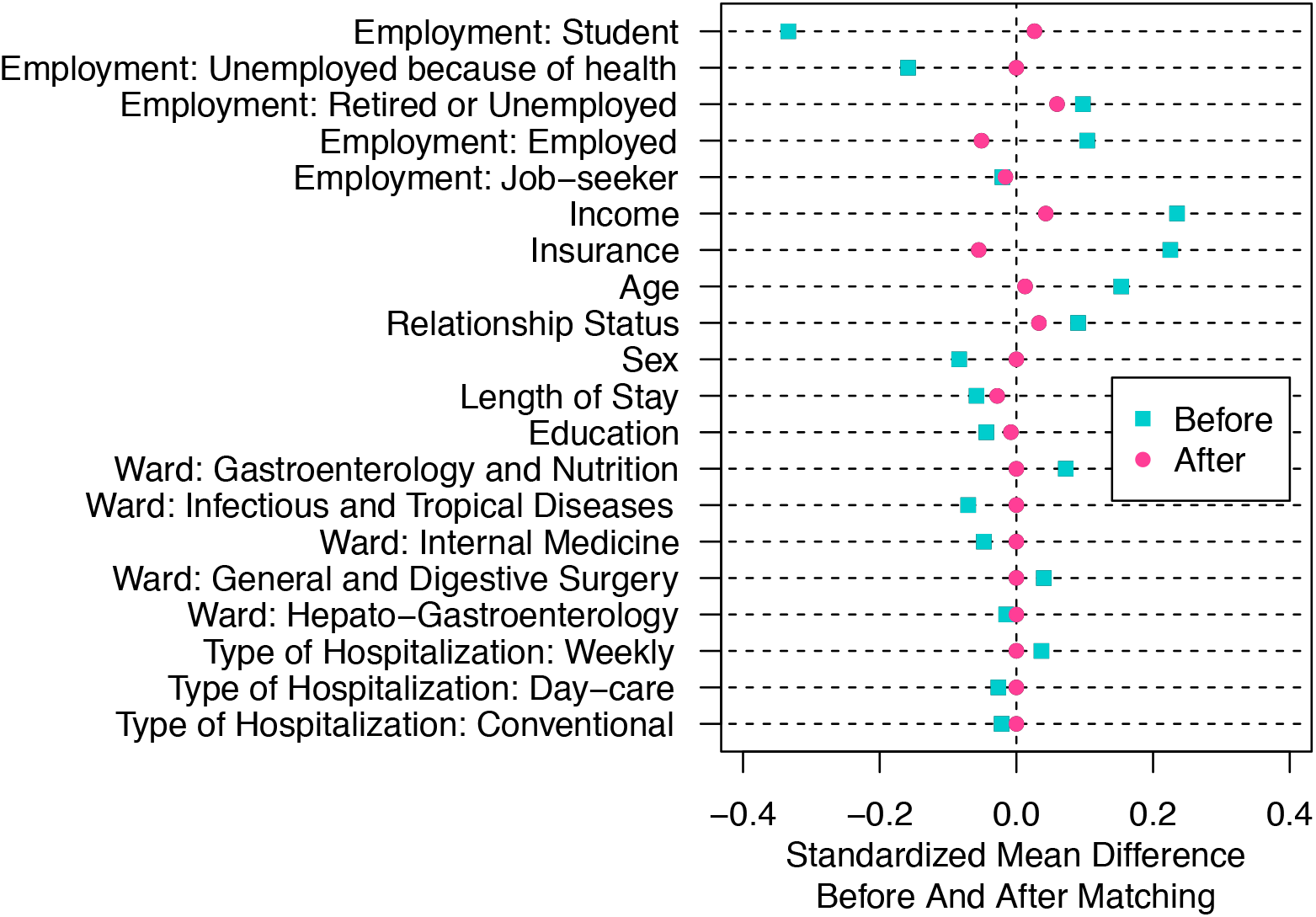
Differences in baseline variables between Internet and Telephone responders, before and after the matching procedure.

Figure 3 shows the differences between I and T groups, before and after matching, for the eight scales and the two summary measures composing SF-36. Figure 3 indicates that the matching procedure had a limited impact on the differences observed between I and T in each of the components of SF-36: regardless of the value of the difference before matching, the corresponding difference after matching appeared similar. Importantly, the means observed in the Telephone group were all above the corresponding values observed in the Internet group.

**Figure 3:**
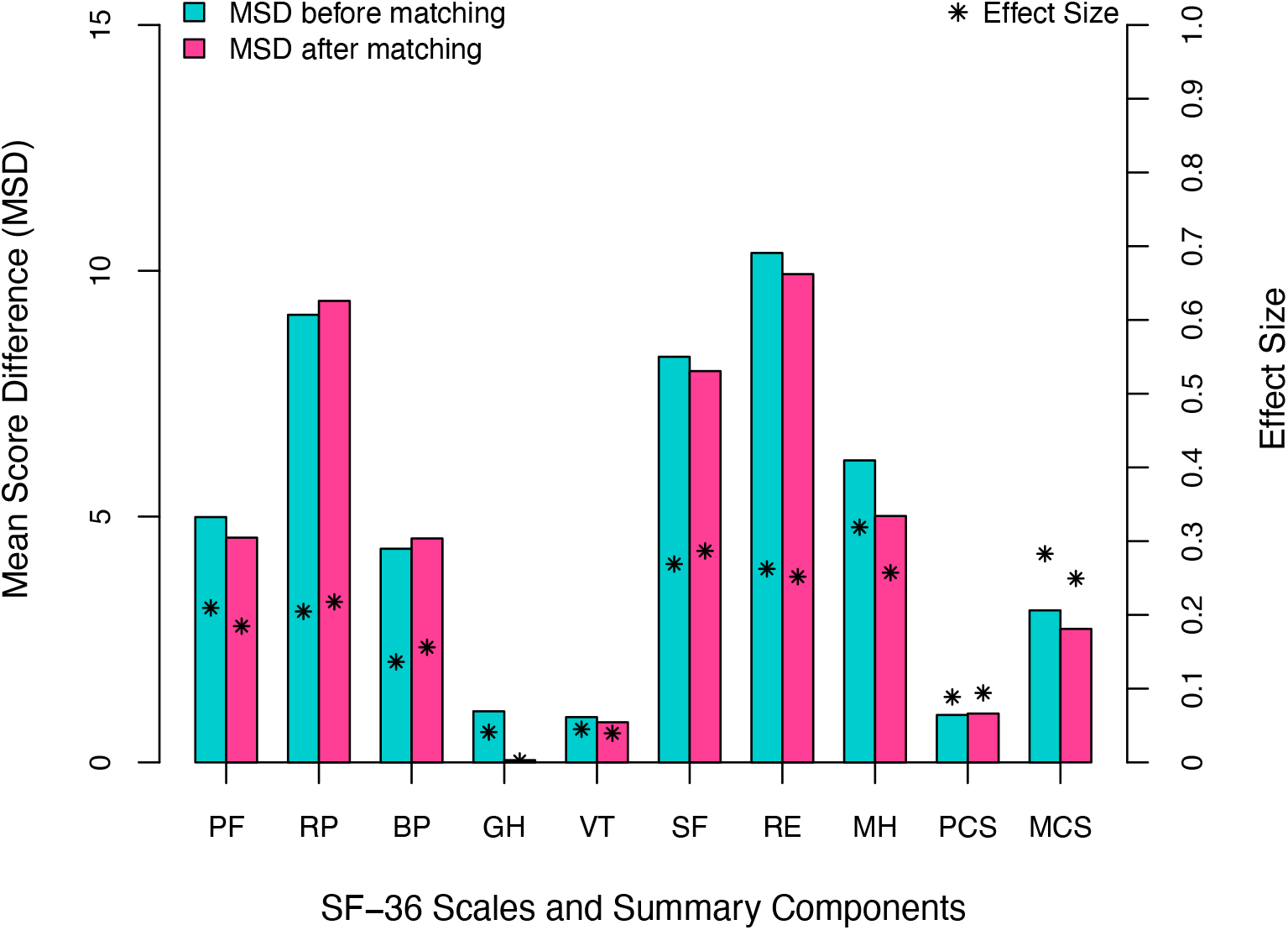
Observed mean score differences (Telephone - Internet) of SF-36 scales and summary components, before and after matching. Abbreviations used: MSD, mean score difference; PF, physical functioning; RP, role-physical; BP, bodily pain; GH, general health; VT, vitality; SF, social functioning; RE, role-emotional; MH, mental health; PCS, physical component summary; MCS, mental component summary.

Table 2 details the results observed after matching. The mean difference between I and T group was greater than five (threshold recommended for a declaring that the difference corresponds to a significant clinical status) for four scales (RP, SF, RE, and MH) with an associated mild effect size ranging from 0.22 to 0.29. Moreover, the difference approached this threshold for two other scales (4.57 and 4.56 for PF and BP, respectively), with a low corresponding effect size, 0.18 and 0.16, respectively. The above-mentioned 6 differences were all statistically significant (see Table 2). In contrast, the observed mean difference between T and I was low for the remaining two scales (0.04 and 0.82 for GH and VT, respectively), and not significant. When examining the physical and the mental component summary, the difference was 0.99 and 2.72, respectively, the latter difference being statistically significant and with an associated effect size at 0.25.

**Table 2:** SF-36 scores in the Internet and Telephone group after matching (n_Internet_ = n_Telephone_ = 245)

| Scale or component summary | Group | Median Score [IQR] | Mean Score [95% CI] | Standard Deviation | Score difference (Telephone – Internet) |  |  |
| --- | --- | --- | --- | --- | --- | --- | --- |
|  |  |  |  |  | P value | Mean Difference | Effect size |
| PF | Internet | 85 [65–95] | 76.08 [72.92–79.08] | 24.56 | 0.012 | 4.57 | 0.18 |
|  | Telephone | 90 [70–100] | 80.65 [77.47–83.71] | 24.93 |  |  |  |
| RP | Internet | 50 [0–100] | 51.53 [46.22–56.73] | 41.67 | 0.002 | 9.39 | 0.22 |
|  | Telephone | 100 [0–100] | 60.92 [55.31–66.43] | 44.59 |  |  |  |
| BP | Internet | 72 [41–100] | 66.84 [63.55–70.11] | 26.12 | 0.045 | 4.56 | 0.16 |
|  | Telephone | 84 [41–100] | 71.40 [67.42–75.40] | 32.23 |  |  |  |
| GH | Internet | 57 [42–72] | 55.10 [52.57–57.65] | 20.47 | 0.999 | 0.04 | 0.00 |
|  | Telephone | 57 [37–77] | 55.15 [51.96–58.34] | 25.90 |  |  |  |
| VT | Internet | 50 [35–65] | 48.29 [45.78–50.80] | 20.16 | 0.566 | 0.82 | 0.04 |
|  | Telephone | 50 [35–65] | 49.10 [46.41–51.78] | 21.30 |  |  |  |
| SF | Internet | 75 [50–100] | 71.17 [68.16–74.18] | 24.27 | <0.001 | 7.96 | 0.29 |
|  | Telephone | 100 [62.5–100] | 79.13 [75.15–82.96] | 31.24 |  |  |  |
| RE | Internet | 100 [33.33–100] | 67.89 [63.13–72.65] | 39.04 | 0.002 | 9.93 | 0.25 |
|  | Telephone | 100 [66.66–100] | 77.82 [72.79–82.59] | 39.84 |  |  |  |
| MH | Internet | 64 [52–80] | 63.56 [61.21–65.91] | 18.77 | 0.002 | 5.01 | 0.26 |
|  | Telephone | 72 [56–84] | 68.57 [65.94–71.10] | 20.20 |  |  |  |
| PCS | Internet | 44.95 [37.27–53.30] | 44.48 [43.20–45.75] | 10.04 | 0.180 | 0.99 | 0.09 |
|  | Telephone | 48.33 [37.81–54.43] | 45.47 [44.09–46.82] | 11.05 |  |  |  |
| MCS | Internet | 47.49 [35.37–52.60] | 44.68 [43.34–46.01] | 10.62 | 0.002 | 2.72 | 0.25 |
|  | Telephone | 50.86 [41.81–55.50] | 47.40 [46.01–48.76] | 11.15 |  |  |  |

Abbreviations used: PF, physical functioning; RP, role-physical; BP, bodily pain; GH, general health; VT, vitality; SF, social functioning; RE, role-emotional; MH, mental health; PCS, physical component summary; MCS, mental component summary; IQR, interquartile range; 95% CI, 95% confidence interval.

## Discussion

To our knowledge, this study is the first reported to date that compared SF-36 questionnaire data collected either via a telephone interview or via a self-completion on a dedicated internet website. The study has additional strengths such as the fact that it is based on a randomized trial, with a substantial number of patients included both arms, a large patient case-mix variability (patients originating from 5 very different hospital wards). The main limitation of the study concerns the selection bias related to respondent status in both arms, but such a bias is inherent to the two corresponding modes of administration, and we tried to mitigate this bias as much as possible by conducting a part of the analyses in a matched sub-population. The detailed analysis comparing the scores observed in the whole set of respondents (before matching) and in a sub-population enhancing the similarity of the individuals compared (after matching) constitutes an important strength of the study.

Despite the reminders sent to the patients, Internet group response rate (29%) to survey was dramatically lower than that of the Telephone group (75%) but still within the range of a meta-analysis on Web-based surveys that reported a median participation rate at 27% [36]. While no study compared telephone and internet administration modes for SF-36, two of the four studies that compared telephone and postal mail (paper) administration resulted in higher participation rate in the paper group [18, 23] and the other two had the opposite result [24, 26]. In addition, the participation rates observed in our study are close to those of Basnov et al [13] who reported a lower response rate in the Internet group than that observed in the paper group (23% versus 76%, respectively). In our view, the response rates observed in a survey involving internet versus another method of administration are difficult to interpret and are not generalizable at all: the modes of administration include underlying elements of the whole survey process for which the impact on participation rate is hardly assessable / describable, such as the internet web site design in terms of its attractiveness or convenience, or the detailed procedure for reaching participants by telephone. For example, the relative high rate of participation in the telephone group observed in this study is likely related to the fact that the schedule of the telephone interview was arranged with each participant at the moment of his/her enrollment and that moreover, up to three calls were tried whenever the participant was not reached at the first phone call.

Nevertheless, with a perspective of a rigorous comparison between SF36 estimates issued from the I and T groups, the difference of response rates between groups observed in this study raised concerns in terms of selection bias associated to the responder status. Indeed, the difference in the SF36 estimates observed in these two groups may be mainly due to two features: first, the difference of the mode of administration of the questionnaire strictly speaking (self-completion of patient via the Internet versus completion of a research technician via a telephone interview with the patient), and second, unbalanced characteristics of the individuals in the two groups issued from a selection bias of the respondents (an unavoidable situation inherent to the modes of administration of the questionnaire). Assessing the respective impact of these two features on the observed differences between the SF-36 scores observed in I and T respondents is of primary importance, and in order to get more insight into this issue, we developed a procedure in which responders of the Internet group were matched to similar responders of the Telephone group, according to their baseline characteristics, and we further examined how the score differences between the two groups changed in this matched sample, as compared to the score differences observed in the initial unmatched populations. Figure 2 shows that the matching procedure highly succeeded for composing a sample of similar match-paired patients, but the very modest impact of this matching procedure on modifying the initial score differences between the scores in I and T groups (see Figure 3) highly suggests that the score differences between I and T are mainly attributable to the mode of administration strictly speaking, with a very minor impact of selection bias issues. However and importantly, the scores in the T group were always higher than those in the I group (Figure 3 and Table 2), likely reflecting another type of bias associated with the telephone interview mode of administration: the interviewer effect. Our results are in agreement with previous studies that reported higher SF-36 scores, when administered by telephone compared to those issued from a mailed paper mode of administration [18, 21, 22, 24-26]. Similarly, Lyons et al [37, 38] reported higher scores issued from a face-to-face interview administration than those issued from a self-completion of the SF-36 questionnaire. Altogether, our results and those of previous studies suggest that as compared to patient’s self-completion, the introduction of an interviewer likely acts as a veil that somehow embellish patient’s QoL reported perception. Internet self-completion avoids any potential bias of responses related to an interviewer effect [39], and patients are more likely to freely express their opinions [40] on websites covering anonymity than through telephone. Therefore, self-completion (internet or paper) should probably be preferred for collecting SF-36 data, since the involvement of a third party appears to artificially increase the scores. In any case, our study indicates that an accurate comparison of different scores requires at least avoiding modes of administration of SF-36 mixing self-completion and interview.

For all but two scales out of eight, the mean difference of scores between the groups was statistically significant and higher than 4.5 points (Table 2), and several comments have to be made about this statement of fact. It is worth to recall that the misinterpretations of P values are very common [41, 42]. A statistically significant score difference is not systematically considered as meaningful by authors [43, 44] and Ware et al had initially proposed a 5 points difference between two SF-36 scores as a threshold value for a clinically and socially relevant difference [33]. In our view, considering effect size is an appropriate approach for examining the relevance of score differences because such a perspective takes into account the variability of the measures and not only a rough mean difference threshold. Interestingly, as shown in Table 2, even if there were substantial mean score differences for the majority of the scales between the two different modes of administration, these differences were all related to a small effect size in eight scales and in two summary components of SF-36 according to effect size index classification proposed by Cohen [34]. Cohen defines the small effect size as “noticeably smaller than medium which represents an effect visible to naked eye of a careful observer but also not so small as to be trivial”. On the one hand, the effect size perspective considerably softens the relevance of the observed differences between I and T groups, and raises concerns about considering a five points mean difference as the main critical element of comparison between two scores. Moreover, such results also indicate that in studies involving a substantially variable population, only very large score mean differences would be considered as meaningful when adopting effect size perspective, highly limiting the presumable usefulness of SF-36 in such studies. On the other hand, some score mean differences observed in our study and most likely attributable to the interviewer effect are not negligible. For example, in patients with chronic C hepatitis, Younossi et al [45] have reported a mean value of RP scale at 74.4 and 79.6 in patients with advanced and none to mild fibrosis, respectively (p = 0.0017). Therefore, the differences for RP scale likely attributable to SF-36 mode of administration observed in the present study (51.5 and 60.9 in group I and T, respectively (p = 0.002), see Table 2) are at least comparable to those attributable to substantial different health states reported in other studies.

## Conclusions

As compared to a mode of administration based on telephone interview, the response rate of volunteer patients communicating their SF-36 data via the Internet was much lower, but our study indicates that a substantial proportion of hospitalized patients volunteered for actively documenting their health data via the Internet. Most of all, the study indicates that the telephone interviewer might be viewed as an intermediate subjective pattern in the collection of patient’s data, resulting in a non-negligible increase of SF-36 scores. Therefore, self-administration of SF-36 should be preferred, including via the Internet which is likely a low-cost method. Importantly, the results of this study also strongly advocate for avoiding the conduction of surveys combining methods of SF-36 administration mixing self-reporting and interviews.

## Supporting information

CONSORT checklist

## Data Availability

The datasets analysed during the current study are available from the corresponding author on reasonable request.

## Supplementary Material

Supplemental Table: Internal reliability of SF-36 in the Internet and Telephone group (pdf document).

CONSORT Checklist.

## Ethics approvals

The SENTIPAT study was approved by the Comité de Protection des Personnes Ile de France IX (decision CPP-IDF IX 12-014, June 12, 2012), by the Comité Consultatif sur le Traitement de l’Information en matière de Recherche dans le domaine de la Santé (decision 12.365, June 20, 2012), and by the Commission Nationale de l’Informatique et des Libertés (decision DR-2012-582, December 12, 2012). According to the French law in force at the time of the study, a formal consent of participants was waived and replaced by the following: patients received full information on their participation in the study and the non opposition of each participant in the study was notified (including date of non opposition declaration) in the SENTIPAT study register.

## Competing interests

All authors have completed the ICMJE uniform disclosure form at www.icmje.org/coi_disclosure.pdf and declare: no support from any organization for the submitted work; no financial relationships with any organizations that might have an interest in the submitted work in the previous three years; no other relationships or activities that could appear to have influenced the submitted work.

## Funding

The Assistance Publique–Hôpitaux de Paris (Département de la Recherche Clinique et du Développement) was the trial sponsor.

The SENTIPAT study was funded by grant AOM09213 K081204 from Programme Hospitalier de Recherche Clinique 2009 (Ministère de la Santé).

The sponsor and the funders had no role in study design, data collection and analysis, decision to publish, or preparation of the manuscript.

## Contributors

GH had full access to all of the raw data in the study and can take responsibility for the integrity of the data and the accuracy of the data analysis. Study conception and design: GH. Data acquisition: GH. Analysis AA and GH. Interpretation of data: AA, FC, and GH. First draft of the article: AA and GH. All authors approved the final version of the manuscript.

## Acknowledgements

The SENTIPAT study group:

1. Scientific Committee: Fabrice Carrat, Bérengère Couturier, Gilles Hejblum, Morgane Le Bail, Alain-Jacques Valleron, and the below-mentioned heads and physicians within the poles and units in which patients were recruited. Personnel of the study group within the poles and units concerned:
2. Heads: Marc Beaussier, Jean-Paul Cabane, Olivier Chazouillères, Jacques Cosnes, Jean-Claude Dussaule, Pierre-Marie Girard, Emmanuel Tiret and Dominique Pateron;
3. Additional corresponding physicians: Laurent Beaugerie, Laurence Fardet, Laurent Fonquernie, François Paye, Laure Surgers.
4. Nursing supervisors: Françoise Cuiller, Catherine Esnouf, Hélène Haure, Valérie Garnier, Josselin Mehal-Birba, Nelly Sallée, and Sylvie Wagener.

The authors are indebted to the excellent technical team of the study:

The clinical research technicians: Élodie Belladame, Azéline Chevance, Magali Girard and Laurence Nicole, who included patients, collected baseline data and interviewed the patients followed-up by telephone.

The software staff, especially Pauline Raballand, but also Frédéric Chau and Frédéric Fotré, who created and maintained the trial’s dedicated website;

We thank all the medical and nursing and administrative staff of the General and Digestive Surgery (including Ambulatory Surgery), Gastroenterology, Hepatology, Infectious Diseases and Internal Medicine departments of Hopital Saint-Antoine;

We thank all patients who participated in the study.

## Notes

### Competing Interest Statement

The authors have declared no competing interest.

### Clinical Trial

ClinicalTrials.gov NCT01769261

### Clinical Protocols

https://clinicaltrials.gov/ct2/show/NCT01769261

### Author Declarations

The SENTIPAT study was approved by the Comité de Protection des Personnes Ile de France IX (decision CPP-IDF IX 12-014, June 12, 2012); by the Comité Consultatif sur le Traitement de l'Information en matière de Recherche dans le domaine de la Santé (decision 12.365, June 20, 2012); and by the Commission Nationale de l'Informatique et des Libertés (decision DR-2012-582, December 12, 2012).

