## Supplemental Table 1 for "Substantial SF-36 score differences according to the mode of administration of the questionnaire: an ancillary study of the SENTIPAT multicenter randomized controlled trial comparing web-based questionnaire self-completion and telephone interview"

| SF-36 Scale | Cronbach's alpha [95% confidence interval] | |
|---|---|---|
| | Internet group | Telephone T |
| PF, physical functioning | 0.919 [0.903–0.933] | 0.898 [0.885–0.909] |
| RP, role-physical | 0.861 [0.830–0.887] | 0.931 [0.922–0.939] |
| BP, bodily pain | 0.906 [0.879–0.927] | 0.933 [0.921–0.943] |
| GH, general health | 0.797 [0.754–0.835] | 0.717 [0.681–0.751] |
| VT, vitality | 0.852 [0.819–0.880] | 0.714 [0.677–0.750] |
| SF, social functioning | 0.879 [0.845–0.906] | 0.943 [0.933–0.951] |
| RE, role-emotional | 0.791 [0.741–0.832] | 0.954 [0.947–0.960] |
| MH, mental health | 0.916 [0.898–0.932] | 0.888 [0.874–0.901] |
